# Protective antibodies and T cell responses to Omicron variant three months after the booster dose of BNT162b2 vaccine

**DOI:** 10.1101/2022.03.04.22271890

**Authors:** Paul Naaber, Liina Tserel, Kadri Kangro, Epp Sepp, Virge Jürjenson, Jaanika Kärner, Liis Haljasmägi, Uku Haljasorg, Marilin Kuusk, Joachim M. Gerhold, Anu Planken, Mart Ustav, Kai Kisand, Pärt Peterson

## Abstract

The high number of mutations in the Omicron variant of SARS-CoV-2 cause its immune escape when compared to the earlier variants of concern (VOC). At least three vaccine doses are required for the induction of Omicron neutralizing antibodies and further reducing the risk for hospitalization. However, most of the studies have focused on the immediate response after the booster vaccination while the duration of immune response is less known. We here studied longitudinal serum samples from the vaccinated individuals up to three months after their third dose of the BNT162b2 vaccine for their capacity to produce protective antibodies and T cell responses to Wuhan and Omicron variants. After the second dose, the antibody levels to the unmutated spike protein were significantly decreased at three months, and only 4% of the individuals were able to inhibit Omicron spike interaction compared to 47%, 38%, and 14% of individuals inhibiting wild-type, delta, and beta variants’ spike protein. Nine months after the second vaccination, the antibody levels were similar to the levels before the first dose and none of the sera inhibited SARS-CoV-2 wild-type or any of the three VOCs. The booster dose remarkably increased antibody levels and their ability to inhibit all variants. Three months after the booster the antibody levels and the inhibition activity were trending lower but still up and not significantly different from their peak values at two weeks after the third dose. Although responsiveness towards mutated spike peptides was lost in less than 20 % of vaccinated individuals, the wild-type spike-specific CD4+ and CD8+ memory T cells were still present at three months after the booster vaccination in the majority of studied individuals. Our data show that two doses of the BNT62b2 vaccine are not sufficient to protect against the Omicron variant, however, the spike-specific antibodies and T cell responses are strongly elicited and well maintained three months after the third vaccination dose.

## Introduction

The Omicron variant of SARS-CoV-2 has caused breakthrough infections worldwide among previously vaccinated individuals showing that the antibody levels formed after the vaccination are not sufficient to inhibit its transmission ^1^. The variant has many new mutations compared to previous variants of concern (VOCs), which has enabled an escape from neutralizing antibodies and added to its transmission efficiency in communities ^2^. Many of the mutations are located in spike protein that is needed for the virus entry to human cells and is targeted by most of the currently available vaccines ^3^.

Despite increased infections, the risk for severe disease and hospitalization with Omicron among vaccinated individuals is reduced compared to earlier variants, including Delta ^4^. As one reason, animal studies have shown that Omicron was limited to the upper respiratory tract causing less damaging infection in the lungs, and these findings have been supported by human ex vivo studies ^5^. Secondly, the high number of mutations in spike protein has added to its ability to escape from neutralizing antibodies and develop resistance to therapeutic antibodies in clinical use ^6-11^. The variant efficiently escapes from the neutralizing antibodies in individuals who have been vaccinated with two doses of mRNA vaccine ^7,10^. However, the third dose of mRNA vaccine has been reported to prominently induce neutralizing antibodies against the Omicron variant inhibiting the loss of neutralizing activities ^10^. Furthermore, the breakthrough infection by non-Omicron variant robustly elicits Omicron-neutralizing antibodies in the vaccinees who have received mRNA vaccines ^12^, suggesting that three contacts with the viral antigen either in form of vaccination or breakthrough infections are sufficient to avoid severe Covid-19. Thus, the hospitalization with the Omicron variant was found 65% and 80% lower for those who received 2 or 3 doses, respectively, when compared to those who had not received any vaccination ^13 14,15^. Apart from the neutralizing antibodies, the vaccination and previous infections with SARS-CoV-2 induce<u>d</u> robust CD4+ and CD8+ T cell responses that largely cross-reacted with Omicron despite its high rate of spike gene mutations ^16-18^.

In addition to the number of vaccinations, another critical factor influencing the immune response to the SARS-CoV-2 is the time since the mRNA vaccinations ^19^. As the antibody levels wane in vaccinated individuals over time, these two factors need to be considered in intraindividual and longitudinal analyses of vaccinated cohorts to identify the antibody and T cell responses elicited after the vaccinations. However, most of the studies have analyzed the immune response shortly after the booster and there is limited data on the durability of the effect of the third dose on Omicron neutralization.

We have followed a cohort of individuals vaccinated with the BNT162b2 vaccine and longitudinally measured their antibody levels and T cell responses to Omicron variant up to three months after the third dose of vaccination. We measured serum capacity to block Omicron spike interaction with ACE2 receptor after each vaccination dose and compared this to previous VOCs. The prevalence of spike-specific CD4+ and CD8+ T cells was studied before and after the booster vaccination and responses towards Omicron mutation peptide pool were compared to the ones elicited by peptides derived from Wuhan spike.

## Results

### Study group

We studied a cohort of diagnostics lab personnel vaccinated with three doses of the BNT162b2 vaccine, which we have earlier followed for their second dose vaccine response ^20^. The details of the study groups are given in **Table 1**. Briefly, the cohort received the first and the second vaccination dose in January 2021, and the third nine months after the start of the trial in October 2021, with their last blood sample collected at the end of January 2022. The collected samples included nine different time points collected before and after the three-dose vaccination; these were before the first dose (B1D), before the second dose (B2D), one week after the second dose (1wA2D), six weeks after the second dose (6wA2D), three months after the second dose (3mA2D), 6 months after the second dose (6mA2D), 9 months after the second dose (9mA2D), two weeks after the third dose (2wA3D) and 3 months after the third dose (3mA3D).

**Table 1.** Summary results at different time-points

|  | Study groups/time-points |  |  |  |  |  |  |  |  |
| --- | --- | --- | --- | --- | --- | --- | --- | --- | --- |
|  | B1D | B2D | 1wA2D | 6wA2D | 3mA2D | 6mA2D | 9mA2D | 2wA3D | 3mA3D |
| **Antibodies to S-RBD** |  |  |  |  |  |  |  |  |  |
| IgG (AU/mL)<br>median/IQR (n) | 1.25/ 0.3-2.5 (88) | 1246/ 666-2582 (111) | 24534/<br>13985-36616<br>(106) | 12752/<br>8225-17348 (89) | 5226/3097-6924 (90) | 1383/893<br>-2463<br>(84) | 739/419-1359 (73) | 32899/179<br>14-46718<br>(60) | 13119/<br>6149-21767 (51) |
| **Inhibition of Spike-ACE2 interaction (relative OD)** |  |  |  |  |  |  |  |  |  |
| SARS-CoV-2 (wild<br>type)<br>median/IQR (n) | 0.97/0.95-0.99 (49) |  | 0.33/0.13-0.46 (49) |  | 0.76/0.64-0.83<br>(49) |  | 0.92/0.86-0.96 (71) | 0.13/0.07-0.33 (56) | 0.55/0.31-0.70 (51) |
| Beta (B.1.351)<br>median/IQR (n) | 1.00/0.97-1.01 (9) |  | 0.64/0.50-0.71 (49) |  | 0.86/0.79-0.91<br>(49) |  | 0.90/0.86-0.96 (71) | 0.23/0.14-0.42 (56) | 0.47/0.26-0.6 (51) |
| Delta (B.1.617.2)<br>Median/IQR (n) | 0.99/0.97-1.02 (49) |  | 0.46/0.29-0.58 (49) |  | 0.80/0.68-0.84 (49) |  | 0.89/0.86-0.95 (71) | 0.16/0.09-0.36 (56) | 0.59/0.41-0.73 (51) |
| Omicron (B.1.1.529)<br>Median/IQR (n) | 0.96/0.92-1.03 (48) |  | 0.79/0.68-0.85 (49) |  | 0.90/0.81-0.97 (49) |  | 0.87/0.82-0.97 (71) | 0.44/0.31-0.58 (56) | 0.64/0.44-0.73 (51) |
| **T cells** |  |  |  |  |  |  |  |  |  |
| Spike-specific CD8+ T<br>cells (% from CD8+,<br>Median/IQR (n)) |  |  |  |  | 0.070/<br>0.008-<br>0.153 (79) |  | 0.045/0-<br>0.24 (68) | 0.31/0.13-<br>0.45 (51) | 0.11/0.04-<br>0.27 (43) |
| Spike-specific CD4+ T<br>cells (% from CD4+,<br>Median/IQR (n)) |  |  |  |  | 0.245/<br>0.008-<br>0.510 (78) |  | 0.185/0.06-<br>0.38 (68) | 0.49/0.3-<br>0.82 (51) | 0.34/0.23-<br>0.61 (43) |
| Omicron-specific CD8+<br>T cells (% from CD8+,<br>Median/IQR (n)) |  |  |  |  |  |  |  |  | 0.06/0.01-<br>0.16 (43) |
| Omicron-specific CD4+<br>T cells (% from CD4+,<br>Median/IQR (n)) |  |  |  |  |  |  |  |  | 0.14/0.06-<br>0.42 (43) |

B1D - before the first dose, B2D - before the second dose, 1wA2D - one week after the second dose, 6wA2D -
six weeks after the second dose, 3mA2D - three months after the second dose, 6mA2D - 6 months after the
second dose, 9mA2D - 9 months after the second dose, 2wA3D - two weeks after the third dose, 3mA3D - 3 months after the third dose

### SARS-CoV-2 antibody dynamics after three-dose vaccination

Before the third vaccination, which was given nine months after the second dose (9mA2D), the S-RBD IgG levels were significantly declined (median 739 AU/mL; IQR 419 – 1359; p<0.0001) compared to the second dose of vaccination (**Figure 1, Table 1**). Two weeks after the third dose (2wA3D) S-RBD IgG levels increased to a median 32899 AU/mL (IQR 17914 – 46718; p<0.0001) as compared to the pre-vaccination (9mA2D) time point. However, 3 months after the third dose (3mA3D), the S-RBD IgG levels were trending lower (median 13119 AU/mL; IQR 6149-21767) but significantly not different from 2wA3D (p= 0.0713). As in earlier timepoints ^20^, the age of vaccinated individuals had a significant negative correlation with S-RBD IgG response before (9mA2D: r= -0.30, p=0.009), and after the third dose (2wA3D: r= -0.31, p=0.015; 3mA2D: r=-0.39, p=0.004).

**Figure 1.**
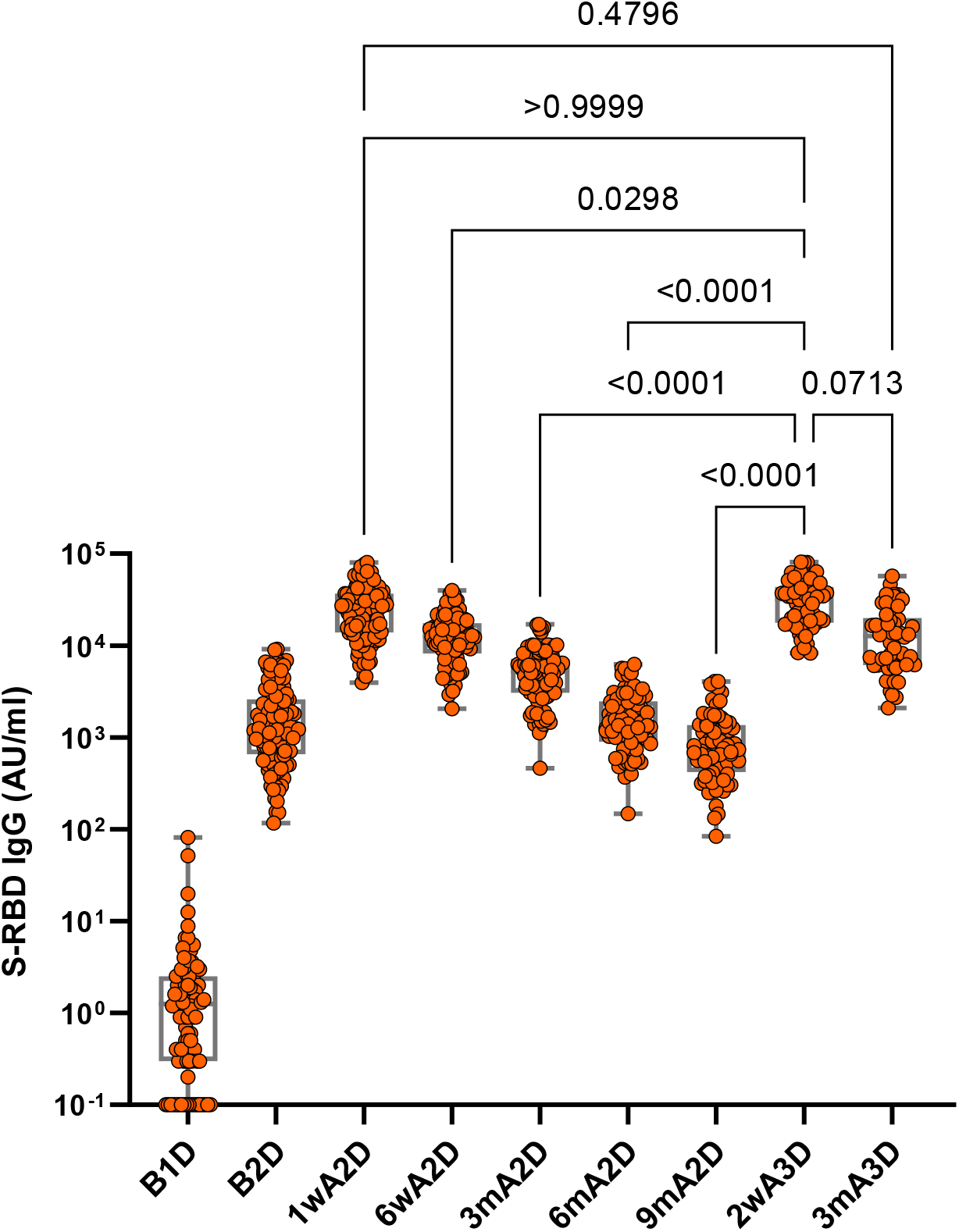
Antibody responses in individuals vaccinated with BNT162b2 vaccine up to three months after the third dose. S-RBD IgG levels before vaccination (B1D), after the single vaccination (B2D); and 1 week (1wA2D); 6 weeks (6wA2D); 3 months (6mA2D); 6 months (6mA2D); 9 months (9mA2D) after the second vaccination dose and 2 weeks (2wA3D) and 3 months (3mA3D) after the third vaccination dose. The dot plot comparisons were performed with the Kruskall-Wallis test and Dunn’s multiple testing correction; p-values >0·0001 are reported as exact numbers.

### Serum inhibition of trimeric spike RBD-ACE2 interaction

We next studied the effectiveness of two and three vaccination doses for their potential to neutralize cell entry of the Omicron variant in comparison to other VOCs. For this, we used an established experimental assay that measures the serum capacity to block the angiotensin-converting enzyme 2 (ACE2) receptor interaction with the SARS-CoV-2 trimeric S protein RBD and thus indicating neutralizing capacity. Using the assay, we tested the longitudinal serum samples from the vaccinated individuals for their inhibition of SARS-CoV-2 Omicron, Delta, Beta, and wild-type spike protein interaction.

Expectedly, the samples from unvaccinated time points did not inhibit the spike-ACE2 interaction of Omicron or any other variant (**Figure 2 and 3, Table 1**). At 1wA2D, most samples were able to block the WT SARS-CoV-2 (98%) and Delta (96%), and slightly less Beta (80%) VOC, however the capacity to inhibit the Omicron variant was significantly less efficient (p<0.0001 compared to all other variants) with only 41% of samples below the inhibition threshold (relative OD 0.75). The blocking capacity decreased over time and was lower for all variants at 3mA2D, most prominently for Omicron (WT 47%, Delta 39%, Beta 14%, and Omicron 4%). Before the third dose, at 9mA2D, the efficiency to block spike binding was further declined with less than 6% of serum samples over the threshold value for all variants, indicating that the time after the second dose had a strong effect on active immune protection but increasingly declined thereafter. After the administration of the third dose at 2wA3D, all samples recovered their blocking activity and were able to inhibit the binding of the spike protein of wild-type, Beta, and Delta VOCs. Nevertheless, 5.5% of the sera did not reach the threshold even after three-dose vaccination when the spike of the Omicron variant was tested (**Figure 2 and 3**). At the last time point, three months after the booster (3mA3D), all SARS-CoV-2 variant spike proteins were inhibited by the majority of immunized sera (>80% of samples), however, the increase in medians (range 0.13 - 0.44 at 2wA3D vs 0.47 - 0.64 at 3mA3D) of the relative OD values for all variants suggested a diminished blocking activity for all variants, consistent with S-RBD IgG results at 3mA3D.

**Figure 2.**
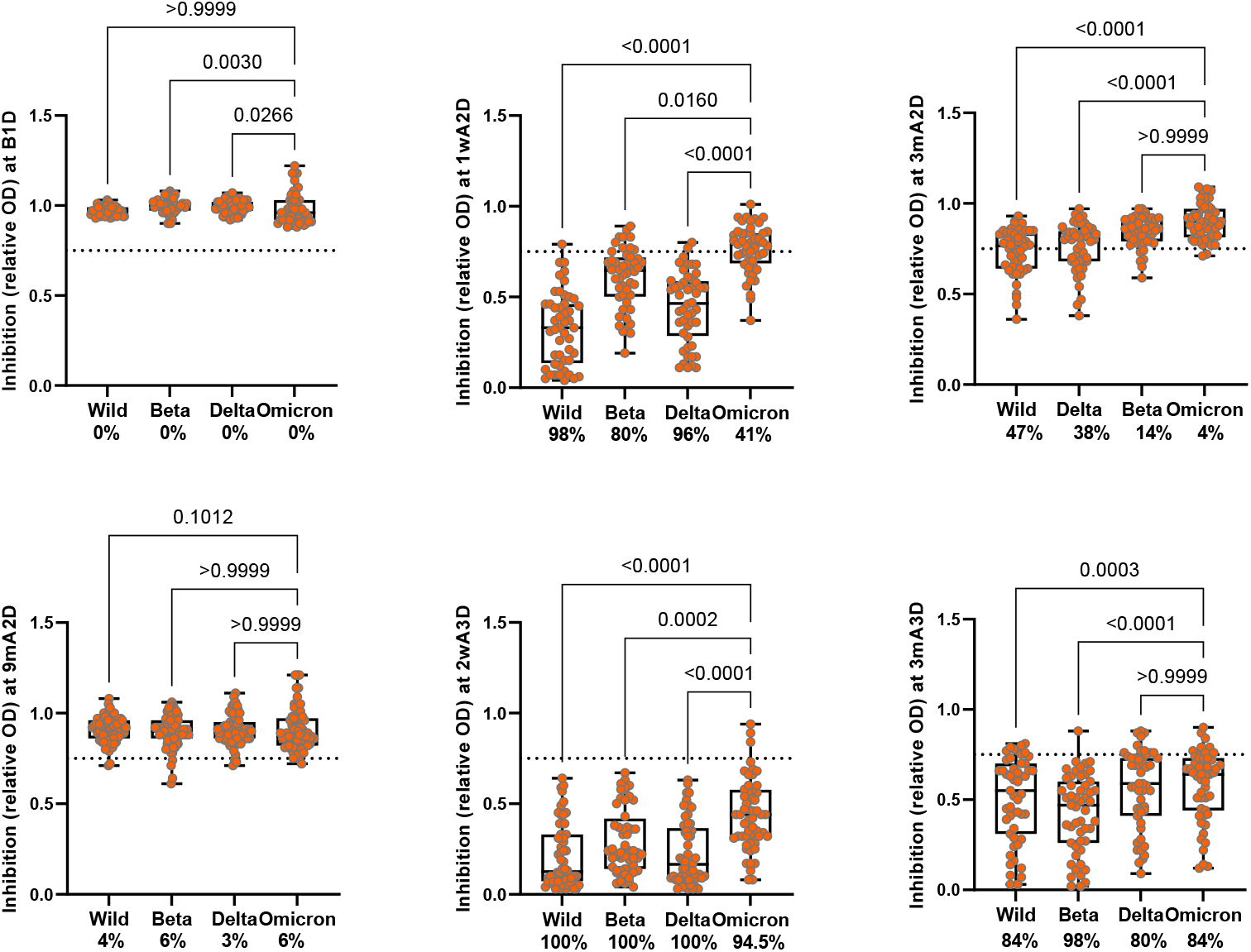
Inhibition of ACE2-trimeric Spike interaction by vaccine-induced antibodies. Serum antibody capacities to block the interaction of ACE2 receptor and Spike protein of wild type, Beta, Delta and Omicron variants before the first dose (B1D), at 1 week (1wA2D), 3 months (3mA2D) and 9 months after the second dose, and 2 weeks (2wA3D) and 3 months (3mA3D) after the third dose of vaccinations. The dotted line indicates the relative OD value of 0.75, which is a threshold for sufficient blocking of ACE2 binding. The matched data analysis was performed with the Friedman test with Dunn’s multiple testing correction; p-values >0.0001 are reported as exact numbers. The percentage of samples that were able to reach the threshold of blocking activity is shown below each graph.

**Figure 3.**
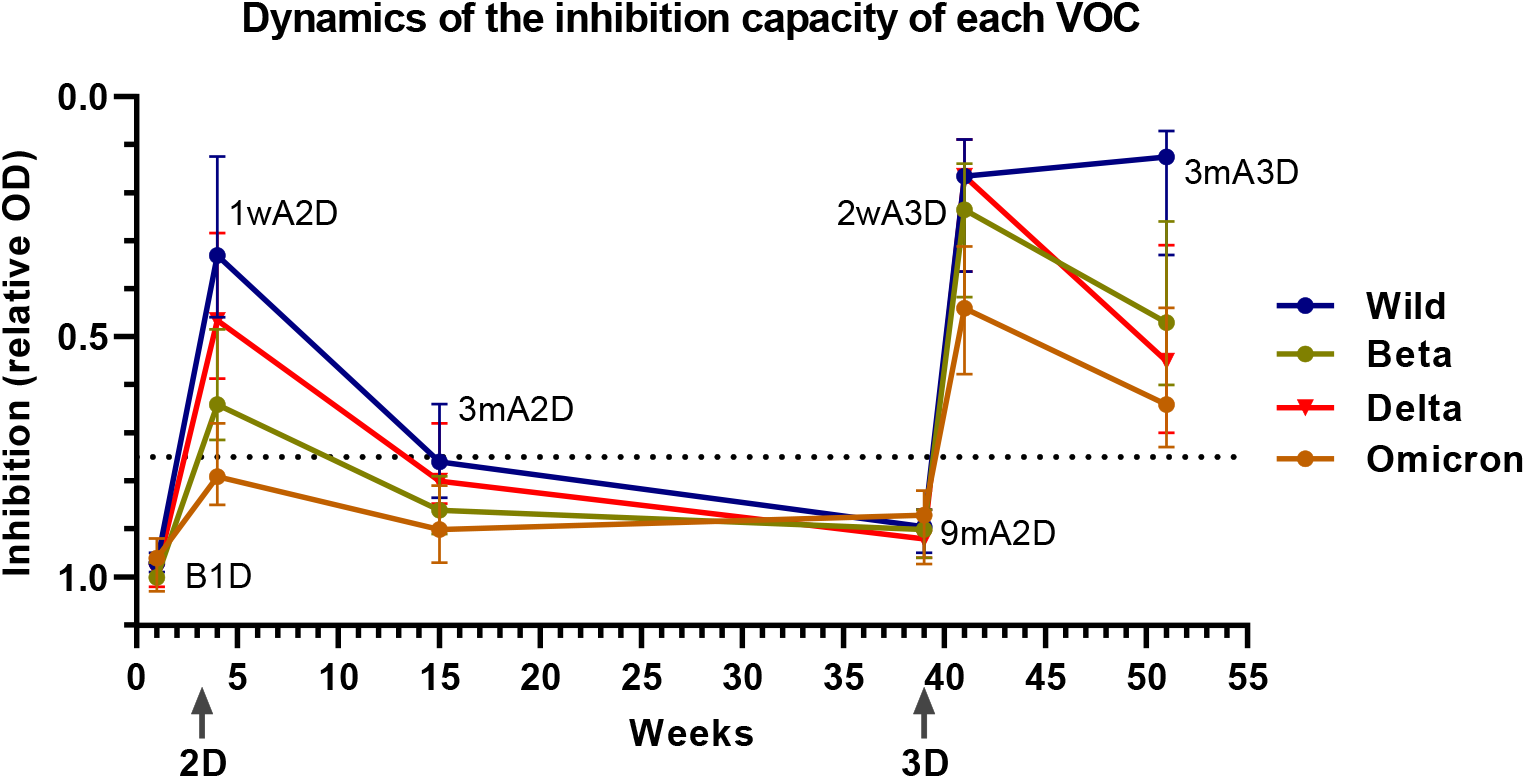
The dynamics of the ACE2 inhibition capacity of wild-type virus, and Beta, Delta and Omicron VOC spike proteins over time. Each line corresponds to one viral strain, the data points show the median values and error bars interquartile ranges of samples collected over time: before the first dose (B1D), 1 week (1wA2D), 3 months (3mA2D), 9 months (9mA2D) after the second dose, 2 weeks (2wA3D) and 3 months (3A3D) after the third dose. The second and third vaccination points are shown at corresponding weeks on X-axis as 2D and 3D, respectively. The relative optical density (OD) values on the inverted Y-axis show the inhibition activity. The value over 0.75 relative OD is considered as the threshold of the serum sample to block ACE2-spike interaction.

Our previous study showed a strong correlation between the S-RBD IgG levels and blocking antibodies ^20^. The Spearman rank correlation analysis at each vaccination point showed that, in contrast to other variants, Omicron had a moderate correlation with S-RBD IgG and other VOCs (r=0.57-0.63) immediately after the second dose (**Figure 4**). However, at subsequent sample collection points at 3mA2D and 9mA2D, we did not find the Omicron results to correlate with S-RBD IgG or the inhibition capacity of other variants indicating a deterioration of the protection against Omicron VOC. Strikingly, the Omicron variant’s correlation with S-RBD IgG and blocking capacity to other variants was restored after the booster dose (r=-0.85 for IgG RBD and r>0.90 for the inhibition experiments) and maintained a very strong correlation even at 3mA3D (r=-0.83 for IgG RBD and r>0.92 for the inhibition experiments). These data strongly support the requirement of three doses to gain protective antibodies against Omicron.

**Figure 4.**
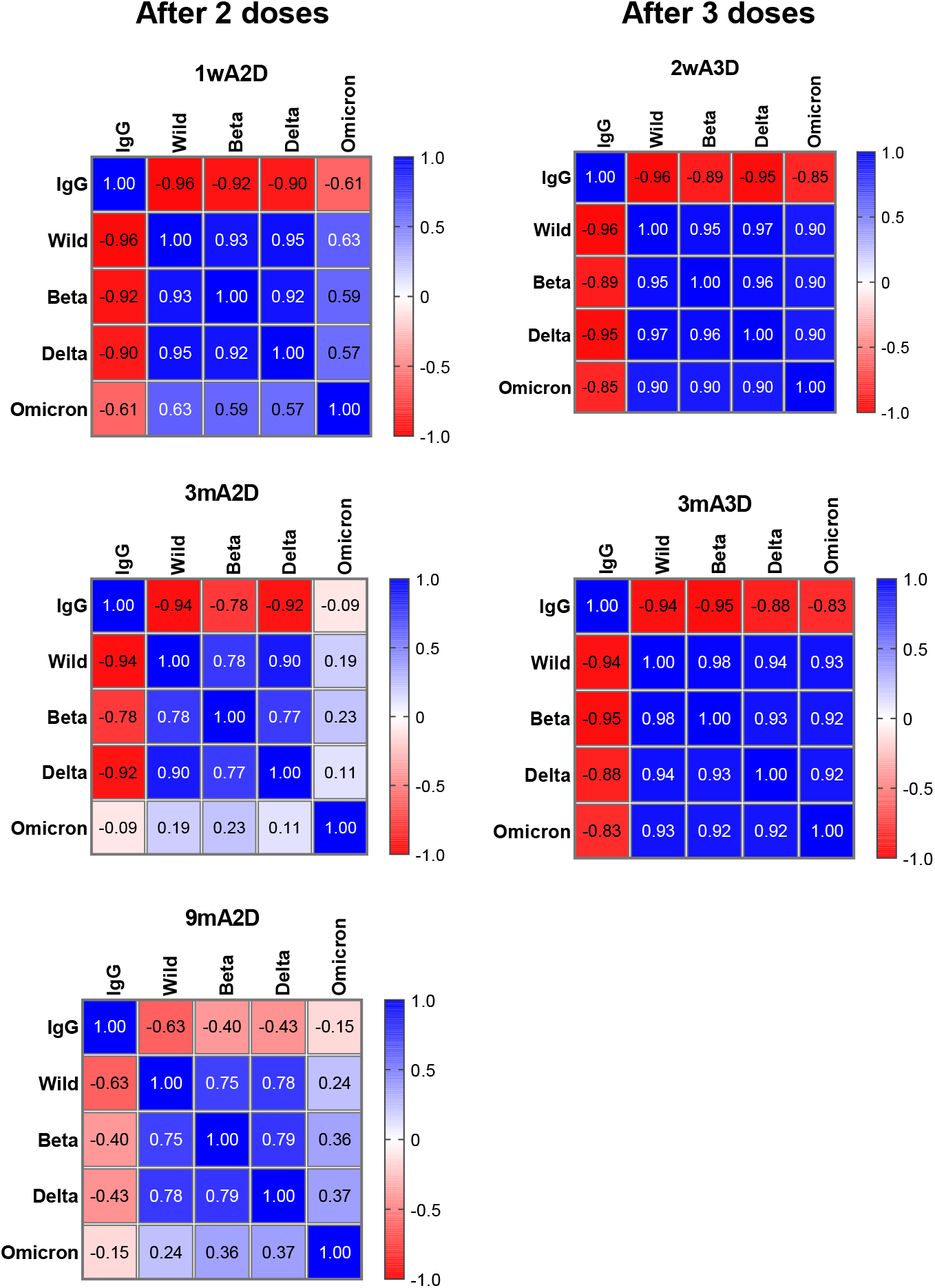
Correlation between serum S-RBD antibody levels and the inhibition capacity of ACE2-spike interactions. Spearman rank correlation analysis at each sample collection time after two and three vaccination doses. Neutralization data is in relative optical density (OD) values with inverted direction (lower values indicate stronger inhibition) and hence they are in negative correlation with S-RBD IgG (blue) and positive correlation (red) between each other.

### T cell responses

Several studies have reported stable or slightly diminished T cell responses to Omicron VOC immediately after the booster dose. We here investigated the percentage of activation-induced marker positive (AIM) memory T cells in CD4+ and CD8+ T cell compartments before and after the third dose, and the Omicron-specific CD4+ and CD8+ T cell responses at three months after the booster dose (3mA3D). We found 84% and 58% of vaccinated individuals to have CD4+ and CD8+ memory responses before the third dose at 9mA2D (**Figure 5**) while the percentage of AIM+ CD4+ or CD8+ T cells was not significantly different from the values measured 6 months earlier at 3mA2D. Administration of the third dose (2wA3D) increased these percentages to 100% and 90%. The frequency of S-specific T cells was significantly higher in 2wA3D group among CD4+ (median/IQR: 0.49/0.3 – 0.82) compared to 9mA2D (0.18/0.06 – 0.38; p<0.0001) and CD8+ T cells (median/IQR: 0.31/0.13 – 0.45 vs 0.04/0 – 0.24; p<0.0001). At 3mA3D 97% of the vaccinees still harboured S-specific CD4+ T cells (median/IQR: 0.34/0.23 – 0.61) and 79% had AIM+ CD8+ T cells (median/IQR: 0.11/0.04 – 0.27) that was not significantly different from the previous time point for CD4+ T cells, but slightly decreased for CD8+ T cells. At 3mA3D AIM+ CD4+ T cell percentage correlated negatively with age (r=-0.38, p=0.01) and positively with S-RBD IgG (r=0.34; p=0.025).

**Figure 5.**
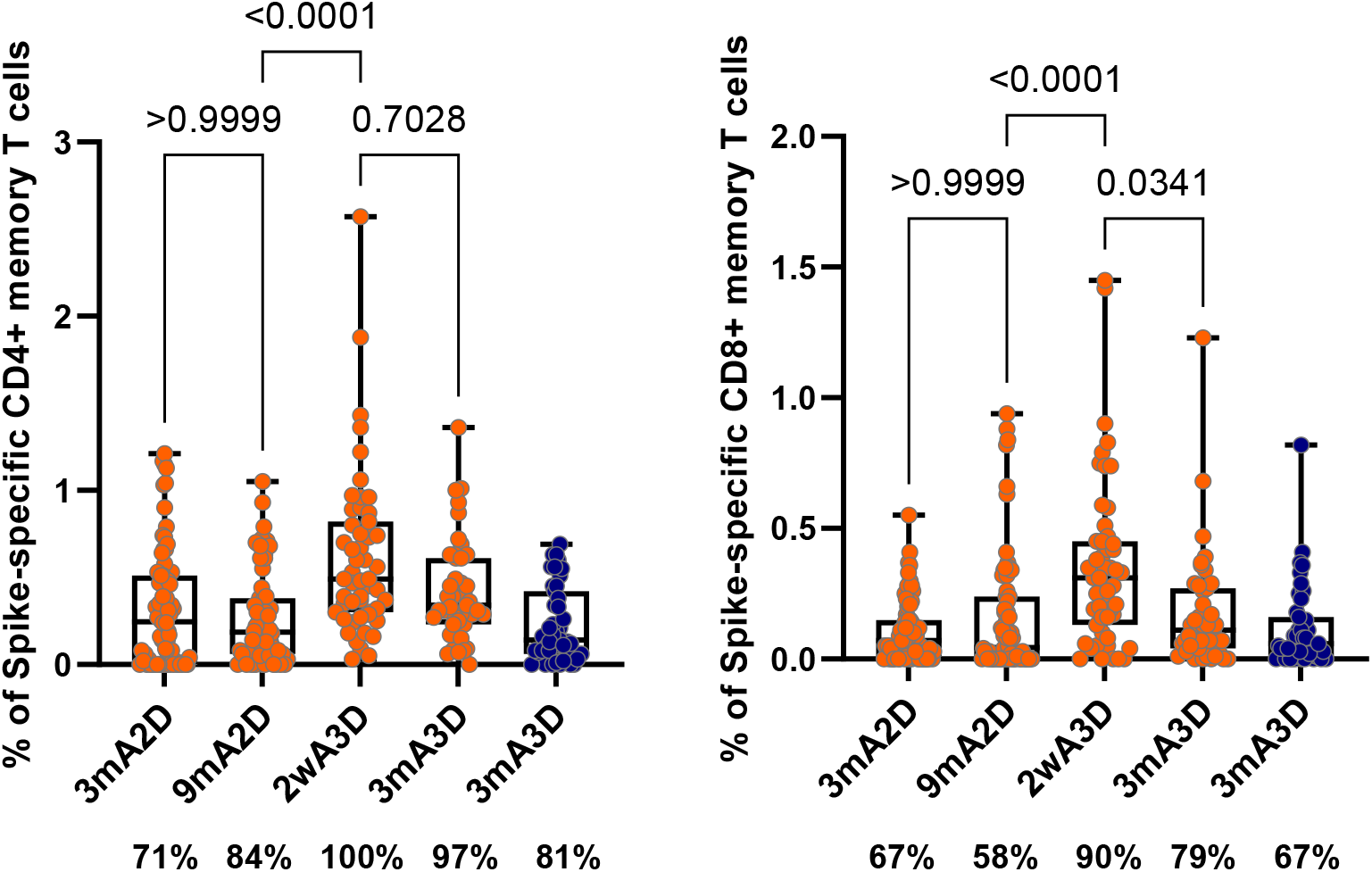
Spike-specific T cell responses in vaccinated individuals. Postvaccination frequency of S-specific (A) CD4^+^ and (B) CD8^+^ T cells at 3 months (3mA2D), 9 months after the second dose (9mA2D), 2 weeks after the third vaccination dose (2wA3D) and 3 months after the third dose (3mA3D). Red dots represent the percentage of T cells that responded to peptide pools derived from Wuhan variant spike sequence and blue dots correspond to the percentage of T cells that responded to peptide pool consisting selectively from Omicron mutated sequences. The data were analysed with the Kruskal-Wallis test with Dunn’s multiple testing correction. The percentage of samples that had spike responsive T cells is shown below each graph.

As the Omicron variant has significantly fewer fully conserved spike epitopes for CD4+ and CD8+ T cells than the previous VOCs ^21^, T cell responses induced by mRNA vaccines coding Wuhan spike, could be also affected. To check this scenario, we performed in parallel T cell stimulation with the peptide pool containing only mutated peptides from Omicron spike (SARS-CoV-2 Prot_S B.1.1.529 Mutation Pool from Miltenyi Biotec) at 3mA3D. We found that only 17% and 15% of the vaccinated individuals with AIM+ T cells towards Wuhan spike, had lost responsiveness against the mutated epitopes by CD4+ and CD8+ T cells respectively.

Collectively, these results showed that the majority of the vaccinated individuals developed efficient SARS-CoV-2 virus-specific memory T cell responses after the second dose, the response was boosted after the third dose, and that both CD4+ and CD8+ memory T cells to the viral antigen persisted at three months after the booster, albeit with slightly lower T cell response against the Omicron variant.

### Predictors of vaccine response

Altogether 97% of participants reported adverse effects after the third dose. The frequency and severity of side effects after the third dose were similar to those after the second dose (total score median/IQR: 7/3 – 12 vs 6/2 – 12; p=0.7). The common side effects after the third dose were pain or swelling at the injection site (in 90%), fatigue (69%), myalgia (51%), malaise (47%), headache (47%) and chills (36%), and resembled those after the second dose.

We found a significant correlation of severity of side effects after the third and the second dose (r= 0·48, p=0·0003). Thus, the individuals with pronounced side effects after the second dose also tended to have these after the third one. The total score of adverse effects after the third dose significantly correlated with the antibody response to S-RBD (r= 0.53, p=0.001), similar to the effect that we previously reported after the second vaccine dose ^20^.

## Discussion

Omicron has demonstrated its immune evasion among vaccinated individuals. We here investigated the dynamics of neutralizing antibodies to Omicron, Beta and Delta VOCs, and original wild-type virus, and studied samples of the vaccinated cohort for their capacity to inhibit spike protein interaction with ACE2. Among the emerged VOCs, the Beta and Omicron variants have a greater ability to escape from neutralizing antibodies and Omicron in particular can escape from the neutralizing antibodies in those who have received two mRNA vaccine doses.

Consistent with previous reports ^2,8,11^, we found decreased capacity to block Omicron in vaccinated serum samples. This was particularly obvious after the second vaccine dose when less than half of the vaccinated sera were able to inhibit the reaction, whereas the Beta variant was blocked by most (80%), and wild-type virus and Delta VOC by more than 95% of the samples. However, the blocking efficiency declined over time and at 9 months after the second dose, all serum samples had lost their blocking activity. Together with the significant waning of the S-RBD binding, this result suggests a remarkable decline of protective antibodies at 9 months after the initial two BNT62b2 vaccine doses. However, two weeks after the booster dose, the serum samples regained their blocking activity, which was in line with the elevated antibody levels to S-RBD.

Given the weaker serum activity generated against Omicron’s spike protein, it was important to determine whether the blocking antibody response stays durable in individuals who received the third dose of mRNA vaccine. For this, we measured the antibodies to S-RBD and VOCs three months after the third dose. After their peak values immediately after the third dose, the antibodies to spike RBD were lower three months later. The decline was not significant, however, and was in their median values almost equal to the levels at 1.5 months after the second dose (median 12752 AU/mL, IQR 8209-7538 for 6wA2D vs median 12729 AU/mL, IQR 628-19883 for 3mA3D). The result indicates that over time the antibodies tend to decline after the booster dose albeit more slowly than after the second dose. Interestingly the antibody blocking efficiency after the third dose remained on the same level for wild-type but was slightly declining with VOCs. Nevertheless, individual serum samples retained a strong correlation between S-RBD levels and the blocking capacity of wild type and the variants, including Omicron. This is in contrast to the inhibition activity that we saw after the second dose, where protection against Omicron did not correlate with S-RBD IgG antibody levels and was discordant from the inhibition of wild-type, Beta, and Delta VOCs. The additional booster dose may elicit Omicron-neutralizing memory B cell subsets and expand the repertoire of the broadly neutralizing antibodies ^22^. In this case, it is plausible that the Omicron-neutralizing clones were acquired as a result of affinity maturation of those with neutralizing activity to previously recognized wild-type spike vaccine antigen.

The waning of antibody levels over time and the emergence of immune escape mutants have turned the focus towards antigen-specific T cells as important players in preventing serious COVID-19 ^23^. The durability of cellular response is one of the key parameters to consider in planning future booster vaccination strategies. mRNA vaccines have shown potent induction of Spike-specific CD4+ and CD8+ responses by the first two vaccine doses but significant contraction of the number of circulating antigen-specific T cells during the first 3 months that stabilize thereafter ^19,24^. This was evident also in our study: the percentages of AIM+ CD4+ and CD8+ T cells were stable between 3 and 9 months after the second dose. However, we demonstrated here that the booster dose of BNT162b2 increased the percentage of both subtypes of Spike-reactive T cells. This is in line with recent reports that have measured immune responses shortly after the booster dose ^25^. Importantly, we found that Spike-specific CD4+ T cell percentages were still elevated 3 months after the third dose of BNT162b2, and AIM+ CD8+ T cells showed only a moderate decline in comparison to 2wA3D time point.

Another important concern about the mRNA vaccine induced T cell response is its cross-reactivity towards VOCs. The Omicron variant has multiple mutations in its spike protein and in comparison to previous VOCs is associated with the fewest fully conserved spike epitopes (72% for CD4+ and 86% for CD8+) ^21^. This means that nevertheless the majority of T cell epitopes are unaffected by the Omicron spike mutations. Indeed, several studies have already confirmed that T-cell reactivity to the Omicron variant is rather well preserved in the majority of vaccinated individuals^16,17,21,25,26^. At 3mA3D we stimulated PBMC samples in parallel with overlapping peptide pools spanning the entire spike protein sequence of the Wuhan strain or a selected pool of peptides mutated in the Omicron spike. It has been predicted that approximately 70% of the mutated Omicron sequences retain HLA class I binding capacity ^21^, e.g. due to conservative substitutions or changes that don’t impact HLA binding. Our findings are in line with these estimations as only less than 20% of the studied individuals had lost responsiveness towards the mutation pool.

In sum, our results suggest increased durability of humoral and cellular immune responses towards SARS-CoV-2 spike protein after the booster dose of BNT62b2 vaccine and confirm the notion of increase in the breadth of antibody response and preservation of T cell responses towards the Omicron variant.

## Material and Methods

### Recruitment, sample, and data collection

The study group and the blood sample collection procedures were reported in our previous study ^20^. Briefly, SYNLAB Estonia employees volunteering to be vaccinated (2 to 3 doses) with COVID-19 mRNA BNT162b2 (Comirnaty Pfizer-BioNTech) vaccine were invited to participate in the study. Starting January 2021, the first two vaccine doses were given three weeks apart, and the third dose was administered nine months after the second dose. The samples were taken before the first dose (B1D), before the second dose (B2D), one week after the second dose (1wA2D), six weeks after the second dose (6wA2D), three months after the second dose (3mA2D), 6 months after the second dose (6mA2D), 9 months after the second dose (9mA2D), two weeks after the third dose (2wA3D) and 3 months after the third dose (3mA3D). The information about the presence of side effects after the third dose was collected as reported previously^20^.

The study has been approved by the Research Ethics Committee of the University of Tartu on February 15, 2021 (No 335/T-21). Participants signed informed consent before the recruitment into the study. The study was performed in accordance with Helsinki Declaration and followed Good Laboratory Practice.

Description of study group is presented previously ^20^. We excluded the participants diagnosed with COVID-19 before or during the study. The number of participants of each time point are presented in **Table 1**.

### Antibody testing

Serum samples were analyzed for the IgG antibodies to SARS-CoV-2 Spike protein receptor-binding domain (S-RBD) IgG using quantitative Abbott SARS-CoV-2 IgG chemiluminescent micro-particle immunoassay (CLIA) on ARCHITECT i2000SR analyzer (Abbott Laboratories) as described previously ^20^.

### ACE2-Spike interaction blocking assay

The serum capacity to block the angiotensin-converting enzyme 2 (ACE2) receptor interaction with SARS-CoV-2 trimeric S protein receptor-binding domain (RBD) was tested using IVD-CE SARS-CoV-2 Neutralizing Antibody ELISA kit (Icosagen) as described previously ^20^. In brief, the ELISA plates covered with SARS-CoV-2 trimeric S proteins of wild-type (wt, Wuhan), Beta (B.1.351), Delta (B.1.617.2), and Omicron (B.1.1.529) VOCs (Icosagen) were incubated with serum samples in a 1/100 dilution and probed with biotinylated ACE2-hFc protein (Icosagen). Streptavidin-Horseradish Peroxidase was used for colorimetric detection and the light absorbance was measured at 450 nm as optical density (OD) values. The OD values of the measured samples were divided by the mean value of the three repeated samples without serum to obtain relative OD values. The samples with relative OD values of <0.75 (i.e. ≥0.75 was determined as the limit of detection) were considered sufficient in blocking ACE2 binding.

### SARS-CoV-2 Spike-specific CD4+ and CD8+ memory T cell responses

For CD4+ and CD8+ T cell response analysis, freshly isolated PBMCs (2×10^6^ cells) were stimulated with overlapping SARS-CoV-2 S peptide pool (1ug/ml, Miltenyi Biotec, 15-mer sequences with 11 amino acid overlap), and with anti-CD28 and anti-CD49d for 20 hours. CEFX peptides (JPT Peptides) were used as a positive control. After the stimulation T cells were stained for CD3 Brilliant Violet 650, CD4 Alexa Fluor 700, CD8 Brilliant Violet 605, CCR7 Alexa Fluor 488, CD45RA APC, CD69 Brilliant Violet 510, OX40 PE-Dazzle (all from Biolegend), and CD137 PE (from Miltenyi Biotech). Antigen-specific cells were gated according to the upregulation of activation-induced markers (AIM) CD137 and CD69 in memory CD8+ T cells and CD137, OX40, and CD69 in memory CD4+ T cells (percentage calculated from total CD8+ or CD4+ cells respectively) as described and shown previously ^20^. The percentage of AIM positive cells in the negative control sample (diluent with costimulatory antibodies) was subtracted from the value from the stimulated sample. The cut-off level for Spike-specific T cell positivity was drawn to 0·02% according to the data from six unvaccinated individuals. 7-AAD was used for the discrimination of dead cells. Flow cytometry was performed using LSRFortessa (BD Biosciences) and the results were analyzed with FCS Express 7 (DeNovo Software). The gating strategy used

### Statistical analyses

GraphPad version 9 was used for statistical analyses and generation of box and whiskers plots and correlation plots. Variables of data (S-RBD IgG, T cell results, and ACE2-Spike interaction inhibition values, age, and the score of side effects) were considered non-normally distributed and are reported as medians and interquartile range (IQR). Kruskal-Wallis test with subsequent Dunn’s multiple comparison testing was used to analyze the S-RBD IgG data and for T cell response data, Friedman test with Dunn’s multiple comparisons for ACE2-Spike interaction inhibition assay results. The correlations between S-RBD IgG values and inhibition results and age or number of side effects were analyzed using Spearman’s correlation with confidence intervals of 95%. For statistical analyses p-values <0.05 were considered to be statistically significant and p-values >0.0001 are reported as exact numbers.

## Data Availability

Data required to reanalyze the data reported in this paper in pseudonymized form is available from the corresponding author upon request.

## Acknowledgments

We thank the participants of the study. The study was supported by The Centre of Excellence for Genomics and Translational Medicine funded by EU European Regional Development Fund (Project No.2014-2020.4.01.15-0012), and the Estonian Research Council grant PRG377, PRG1117, Icosagen Cell Factory, and SYNLAB Estonia.

## Declaration of interests

The authors have nothing to disclose.

## Notes

### Competing Interest Statement

The authors have declared no competing interest.

## References

1. Viana, R., Moyo, S., Amoako, D.G., Tegally, H., Scheepers, C., Althaus, C.L., Anyaneji, U.J., Bester, P.A., Boni, M.F., Chand, M., et al. (2022). Rapid epidemic expansion of the SARS-CoV-2 Omicron variant in southern Africa. Nature. 10.1038/s41586-022-04411-y.

2. Dejnirattisai, W., Huo, J., Zhou, D., Zahradník, J., Supasa, P., Liu, C., Duyvesteyn, H.M.E., Ginn, H.M., Mentzer, A.J., Tuekprakhon, A., et al. (2022). SARS-CoV-2 Omicron-B.1.1.529 leads to widespread escape from neutralizing antibody responses. Cell 185, 467–484.e415. 10.1016/j.cell.2021.12.046.

3. Martin, D.P., Lytras, S., Lucaci, A.G., Maier, W., Grüning, B., Shank, S.D., Weaver, S., MacLean, O.A., Orton, R.J., Lemey, P., et al. (2022). Selection analysis identifies unusual clustered mutational changes in Omicron lineage BA.1 that likely impact Spike function. bioRxiv. 10.1101/2022.01.14.476382.

4. Veneti, L., Bøås, H., Bråthen Kristoffersen, A., Stålcrantz, J., Bragstad, K., Hungnes, O., Storm, M.L., Aasand, N., Rø, G., Starrfelt, J., et al. (2022). Reduced risk of hospitalisation among reported COVID-19 cases infected with the SARS-CoV-2 Omicron BA.1 variant compared with the Delta variant, Norway, December 2021 to January 2022. Euro Surveill 27. 10.2807/1560-7917.ES.2022.27.4.2200077.

5. Hui, K.P.Y., Ho, J.C.W., Cheung, M.C., Ng, K.C., Ching, R.H.H., Lai, K.L., Kam, T.T., Gu, H., Sit, K.Y., Hsin, M.K.Y., et al. (2022). SARS-CoV-2 Omicron variant replication in human bronchus and lung ex vivo. Nature. 10.1038/s41586-022-04479-6.

6. Liu, Y., Liu, J., Xia, H., Zhang, X., Zou, J., Fontes-Garfias, C.R., Weaver, S.C., Swanson, K.A., Cai, H., Sarkar, R., et al. (2021). BNT162b2-Elicited Neutralization against New SARS-CoV-2 Spike Variants. N Engl J Med. 10.1056/NEJMc2106083.

7. Cameroni, E., Bowen, J.E., Rosen, L.E., Saliba, C., Zepeda, S.K., Culap, K., Pinto, D., VanBlargan, L.A., De Marco, A., di Iulio, J., et al. (2021). Broadly neutralizing antibodies overcome SARS-CoV-2 Omicron antigenic shift. Nature. 10.1038/s41586-021-04386-2.

8. Cele, S., Jackson, L., Khoury, D.S., Khan, K., Moyo-Gwete, T., Tegally, H., San, J.E., Cromer, D., Scheepers, C., Amoako, D.G., et al. (2021). Omicron extensively but incompletely escapes Pfizer BNT162b2 neutralization. Nature. 10.1038/s41586-021-04387-1.

9. VanBlargan, L.A., Errico, J.M., Halfmann, P.J., Zost, S.J., Crowe, J.E., Purcell, L.A., Kawaoka, Y., Corti, D., Fremont, D.H., and Diamond, M.S. (2022). An infectious SARS-CoV-2 B.1.1.529 Omicron virus escapes neutralization by therapeutic monoclonal antibodies. Nat Med. 10.1038/s41591-021-01678-y.

10. Garcia-Beltran, W.F., St Denis, K.J., Hoelzemer, A., Lam, E.C., Nitido, A.D., Sheehan, M.L., Berrios, C., Ofoman, O., Chang, C.C., Hauser, B.M., et al. (2022). mRNA-based COVID-19 vaccine boosters induce neutralizing immunity against SARS-CoV-2 Omicron variant. Cell 185, 457–466.e454. 10.1016/j.cell.2021.12.033.

11. Carreño, J.M., Alshammary, H., Tcheou, J., Singh, G., Raskin, A., Kawabata, H., Sominsky, L., Clark, J., Adelsberg, D.C., Bielak, D., et al. (2021). Activity of convalescent and vaccine serum against SARS-CoV-2 Omicron. Nature. 10.1038/s41586-022-04399-5.

12. Rössler, A., Riepler, L., Bante, D., von Laer, D., and Kimpel, J. (2022). SARS-CoV-2 Omicron Variant Neutralization in Serum from Vaccinated and Convalescent Persons. N Engl J Med. 10.1056/NEJMc2119236.

13. UKHSA (2021). SARS-CoV-2 variants of concern and variants under investigation in England Technical briefing: Update on hospitalisation and vaccine effectiveness for Omicron VOC-21NOV-01 (B.1.1.529). In D. UK Health Security Agency, and o.H.a.S. Care, eds. UK Health Security Agency.

14. Wratil, P.R., Stern, M., Priller, A., Willmann, A., Almanzar, G., Vogel, E., Feuerherd, M., Cheng, C.C., Yazici, S., Christa, C., et al. (2022). Three exposures to the spike protein of SARS-CoV-2 by either infection or vaccination elicit superior neutralizing immunity to all variants of concern. Nat Med. 10.1038/s41591-022-01715-4.

15. Wolter, N., Jassat, W., Walaza, S., Welch, R., Moultrie, H., Groome, M., Amoako, D.G., Everatt, J., Bhiman, J.N., Scheepers, C., et al. (2022). Early assessment of the clinical severity of the SARS-CoV-2 omicron variant in South Africa: a data linkage study. Lancet 399, 437–446. 10.1016/S0140-6736(22)00017-4.

16. Keeton, R., Tincho, M.B., Ngomti, A., Baguma, R., Benede, N., Suzuki, A., Khan, K., Cele, S., Bernstein, M., Karim, F., et al. (2022). T cell responses to SARS-CoV-2 spike cross-recognize Omicron. Nature. 10.1038/s41586-022-04460-3.

17. Gao, Y., Cai, C., Grifoni, A., Müller, T.R., Niessl, J., Olofsson, A., Humbert, M., Hansson, L., Österborg, A., Bergman, P., et al. (2022). Ancestral SARS-CoV-2-specific T cells cross-recognize the Omicron variant. Nat Med. 10.1038/s41591-022-01700-x.

18. Liu, J., Chandrashekar, A., Sellers, D., Barrett, J., Jacob-Dolan, C., Lifton, M., McMahan, K., Sciacca, M., VanWyk, H., Wu, C., et al. (2022). Vaccines Elicit Highly Conserved Cellular Immunity to SARS-CoV-2 Omicron. Nature. 10.1038/s41586-022-04465-y.

19. Goel, R.R., Painter, M.M., Apostolidis, S.A., Mathew, D., Meng, W., Rosenfeld, A.M., Lundgreen, K.A., Reynaldi, A., Khoury, D.S., Pattekar, A., et al. (2021). mRNA vaccines induce durable immune memory to SARS-CoV-2 and variants of concern. 10.1126/science.abm0829.

20. Naaber, P., Tserel, L., Kangro, K., Sepp, E., Jürjenson, V., Adamson, A., Haljasmägi, L., Rumm, A.P., Maruste, R., Kärner, J., et al. (2021). Dynamics of antibody response to BNT162b2 vaccine after six months: a longitudinal prospective study. Lancet Reg Health Eur 10, 100208. 10.1016/j.lanepe.2021.100208.

21. Tarke, A., Coelho, C.H., Zhang, Z., Dan, J.M., Yu, E.D., Methot, N., Bloom, N.I., Goodwin, B., Phillips, E., Mallal, S., et al. (2022). SARS-CoV-2 vaccination induces immunological T cell memory able to cross-recognize variants from Alpha to Omicron. Cell. 10.1016/j.cell.2022.01.015.

22. Kotaki, R., Adachi, Y., Moriyama, S., Onodera, T., Fukushi, S., Nagakura, T., Tonouchi, K., Terahara, K., Sun, L., Takano, T., et al. (2022). SARS-CoV-2 Omicron-neutralizing memory B-cells are elicited by two doses of BNT162b2 mRNA vaccine. Sci Immunol, eabn8590. 10.1126/sciimmunol.abn8590.

23. Sette, A., and Crotty, S. (2021). Adaptive immunity to SARS-CoV-2 and COVID-19. Cell 184, 861–880. 10.1016/j.cell.2021.01.007.

24. GeurtsvanKessel, C.H., Geers, D., Schmitz, K.S., Mykytyn, A.Z., Lamers, M.M., Bogers, S., Scherbeijn, S., Gommers, L., Sablerolles, R.S.G., Nieuwkoop, N.N., et al. (2022). Divergent SARS CoV-2 Omicron-reactive T-and B cell responses in COVID-19 vaccine recipients. Sci Immunol, eabo2202. 10.1126/sciimmunol.abo2202.

25. Liu, Y., Zeng, Q., Deng, C., Li, M., Li, L., Liu, D., Liu, M., Ruan, X., Mei, J., Mo, R., et al. (2022). Robust induction of B cell and T cell responses by a third dose of inactivated SARS-CoV-2 vaccine. Cell Discov 8, 10. 10.1038/s41421-022-00373-7.

26. Naranbhai, V., Nathan, A., Kaseke, C., Berrios, C., Khatri, A., Choi, S., Getz, M.A., Tano-Menka, R., Ofoman, O., Gayton, A., et al. (2022). T cell reactivity to the SARS-CoV-2 Omicron variant is preserved in most but not all individuals. Cell. 10.1016/j.cell.2022.01.029.

